# Suppression of influenza virus infection by rhinovirus interference at the population, individual and cellular levels

**DOI:** 10.1101/2021.08.09.21256656

**Authors:** Kin P Tao, Marc Chong, Jason CS Pun, Joseph GS Tsun, Samuel MW Chow, Calvin SH Ng, Maggie HT Wang, Zigui Chan, Paul KS Chan, Albert M Li, Renee WY Chan

## Abstract

**Background:** Investigations of the natural viral interference effect between rhinovirus (RV) and influenza virus (IV) were conducted in temperate regions. We conducted an epidemiological study in Hong Kong, a major epicentre of influenza virus in the sub-tropical region. RV is the most prevalent respiratory virus year-round and causes asymptomatic to mild symptoms while IV infection exerts a great burden of public health. We aimed to examine the correlation of RV prevalence against IV activity.

**Methods:** Nasopharyngeal aspirates (NPA) collected from patients hospitalized in the regional hospitals from 2015 to 2019 were examined for the presence of respiratory viruses. The correlation of the monthly prevalence between all pairs of virus infection, the co-infection rate and the temporal interference of RV and IV were tested. The viral interference was validated *in vitro* by conducting sequential RV and IV infection in the well-differentiated primary human airway epithelial cells.

**Findings:** A total of 112,926 NPA were evaluated, and the Enterovirus/RV was the most prevalent respiratory virus detected. The negative correlation between EV/RV and IVs prevalence was independent of age and meteorological factors. Co-infection of EV/RV and IV was significantly less when compared with other virus pairs. Prior exposure to RV inhibited the replication of influenza A, B and oseltamivir-resistance stain *in vitro* and the inhibition is replication dependent.

**Interpretation:** Epidemiological surveillance and the sequential infection *in vitro* suggested viral interference between EV/RV and IV operated at the population, individual and cellular levels.

**Funding:** This study was supported by the General Research Fund (Ref: 24107017 and 14103119 to RWYC), Health and Medical Research Fund (Ref: COVID190112 to RWYC) and the Chinese University Direct Grant for Research (Ref: 2019.073 to RWYC).

## Introduction

Influenza virus (IV) confers substantial morbidity and mortality worldwide every year. The existence of natural reservoirs of IV makes it impossible to be eradicated in humans.

Influenza vaccine is the classic preventive measure to attenuate disease severity. However, the viral antigenic drift may limit the duration of vaccine effectiveness,^1^ while the antigenic shift may result in the emergence of new strains.^2^ Antivirals is another prophylactic and therapeutic option with a limited effective time frame.^3^ Moreover, antiviral resistant IV strains have been reported sporadically, with an increasing number of oseltamivir-resistant influenza A (R-IAV) is being found.^4^ Whether or not there is natural viral interference in counteracting the influence of IV in the human population or individual levels is an interesting question to address. Identifying novel interfering factors that confer temporal immunity against pan-influenza and other virulent viral infections would provide alternative options for disease prevention and treatment.

Rhinovirus (RV), on the other hand, is the most frequent respiratory pathogen being detected throughout the year.^5^ However, it catches less attention in the public health aspect as it generally causes mild and self-limiting symptoms, and sometimes it is asymptomatic in healthy individuals.^6^ Nevertheless, infants could have up to six to eight RV infections per year, while adults could have around two to four episodes annually.^7^ Having RV infections repeatedly seems to be unavoidable as it comprises more than 160 distinct genotypes. Among genotypes, they do not confer substantial immunity against each other.^8^ Intriguingly, though the prevalence of RV is high, the epidemiology study on RV is uncommon due to its high genotypic diversity. The enterovirus/rhinovirus (EV/RV) test was not included as a standard test in clinical labs until recent years. In Hong Kong, it has become available in public hospitals since September 2015.

With the advancement for the co-detection of viruses through multiplex PCR, multiple population-wide surveillance programs suggested that seasonality of respiratory viral infection is not only contributed by meteorological factors but also the biological interactions among different viruses. The concept of viral interference, a phenomenon in which a primary virus infection could transiently prevent or inhibit the secondary superinfecting virus,^9^ may also play an important role in influencing the pattern of virus outbreaks. Multiple epidemiological analyses, including studies performed in the United Kingdom and the United States, identified a negative interaction between the prevalence of IVs and RVs,^10,11^ and viral interactions operate at multiple levels.^12^ In the epidemiological part of this study, we investigated correlation of virus prevalence and the temporal interference between RV and IV at the population level and the chance of co-infection at individual level. We validated this concept further by conducting sequential infection study in the differentiated primary human nasopharyngeal epithelial cells (HNEC) and human bronchial epithelial cells (HBEC) to elucidate the inhibitory effect *in vitro*.

## Methods

### Study population and dataset

Nasopharyngeal aspirates (NPA) were collected from patients admitted to the six hospitals under the Hong Kong New Territories East Cluster and were screened routinely for respiratory viruses from September 2015 to December 2019. Multiplex real-time PCR were performed by Public Health Laboratory Centre, Centre for Health Protection, Department of Health, Hong Kong. The respiratory virus panel, including influenza A virus (IAV, with subtyping of H1 and H3), influenza B virus (IBV), influenza C (ICV), parainfluenza viruses 1-4 (PIVs), enterovirus/rhinovirus (EV/RV), respiratory syncytial viruses (RSV) and adenovirus (ADV). The test-negative samples were retained as part of the essential denominator to reflect the prevalence in the community to address the fluctuation in sample size over the study period. This virological data covered nine episodes of IV peaks of four consecutive years. Samples with missing or uncertain entries were excluded (<1%). This study was approved by the Joint Chinese University of Hong Kong – New Territories East Cluster Clinical Research Ethics Committee (CREC: 2015.097 and 2019.120).

### Meteorological data

We obtained the meteorological data including ambient temperature (°C) and mean relative humidity (%) measured at the central monitoring station run by the Hong Kong Observatory. The weekly averages of meteorological record were matched with the prevalence over the study period. As absolute humidity was showed to be associated with the respiratory infections,^13-15^ we employed actual vapour pressure (hPa) as a proxy of this humidity measure. The derivation of actual vapour pressure was based on Teten’s formula,^16^

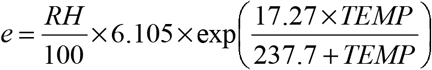

where e, TEMP, and RH denote actual vapour pressure, ambient temperature, and relative humidity respectively.

### Cell lines

H1-HeLa (CRL-1958) and Madin-Darby canine kidney cells (MDCK, CCL-34) were purchased from American Type Culture Collection. Cells were cultured in minimal essential media with non-essential amino acids, 2mM L-glutamine supplemented with 1% penicillin and streptomycin, and 10% fetal bovine serum. Both cell lines were maintained at 37°C and used for RV and IV virus propagation and titration.

### Virus preparation

RV-1B (VR-1645) and RV-A16 (VR-283PQ) were purchased from ATCC and were propagated in H1-HeLa. RV-A16 is a major group RV utilizing ICAM1 as the cellular receptor, and RV-1B belongs to the minor group utilizing LDLR as the cellular receptor, were chosen for this proof-of-concept *in vitro* infection. Seasonal IAV (H1N1 human isolate A/Oklahoma/447/2008), IBV (B/Hong Kong/CUHK/12v033261/2012) and an oseltamivir-resistant H1N1 swine IAV (A/Hong Kong/CUHK/09v071923/2009) were isolated from the NPA of patients and were propagated using MDCK cells. Virus progeny was harvested when the cytopathic effect (CPE) was observed. Virus titers were determined by a viral titration assay.

### Virus titration

H1-HeLa and MDCK cells were seeded on 96-well tissue culture plates one day before the viral titration assay. Cells were washed once with PBS. Virus samples or culture supernatants were titrated in serial half-log_10_ dilutions with the corresponding culture medium before adding the diluted virus to the cell plates in quadruplicate. The highest viral dilution leading to CPE was recorded and the 50% tissue culture infectious dose (TCID_50_) was calculated using the Karber method. The infectivity of RV and IV was monitored by the infectious viral load in the supernatant, as quantitated by viral titration in H1-HeLa or MDCK cells, respectively.

### Airway epithelial cell isolation and differentiation

Human nasopharyngeal epithelial cells (HNEC) and bronchial epithelial cells (HBEC) were isolated from nasopharyngeal flocked swab and bronchial tissue as described ^17^. Briefly, cells were seeded in a 6 well plate coated with human collagen IV and cultured with BEpiCM (Sciencecell): Advanced DMEM (Life Technologies) 1:1 supplemented with HEPES, PS, primocin, glutamax, B27, hydrocortisone, triiodothyronine (T3), epinephrine, N-acetyl-cysteine, nicotinamide, TGFβ inhibitor, BMPi, Rocki, FGF10, FGF7, IGF-1, BSA and R-spondin 1 conditioned medium. Upon confluence, cells were dissociated by TrypLE and seeded onto PureCol coated transwell with a cell density of 2×10^5^cell/well in a 24-well format, and cultured in an air-liquid interface (ALI) using differentiated PneumoCult ALI medium (StemCell) for at least 28 days before infection experiments. Demographics of the donors were provided in **Supplementary Table 1.**

### Experimental design of the sequential RV-A16 and IV virus infection in vitro

Primary HBECs and HNECs were washed with 125ul of PBS five times before infection. Cells were exposed to infection regime 1) RV, 2) IV or 3) a prior RV infection then a IV infection at 48 hpi of the initial RV inoculation at a multiplicity of infection (MOI) of 0.01 (**Figure 6A**). In the infection step, 100ul of RV or sham inoculum were added and allowed for virus attachment for 2 hours at 37°C. The inoculum was discarded, and the cells were washed with PBS twice and the basolateral compartment was replenished with 600ul of the medium. At 48 hours post infection (hpi) of RV, cells were washed with PBS twice before secondary infection of IAVs. The supernatant from the apical compartment of the transwell inserts were collected at 2, 24 and 48 hpi for viral titers determination and gene expression analysis.

### RNA extraction and gene expression analysis

QIAGEN Viral RNA extraction kit and RNaeasy Kit were used according to the manufacturer’s instructions for RNA extraction followed by qPCR. Viral RNA and total RNA were reverse transcribed into cDNA with a PrimerScript RT reagent Kit (Takara). mRNA expression was measured by real-time PCR amplification with SYBR Premix Ex Taq II (Tli RNase H Plus) (Takara) and an ABI Quant Studio 12K real-time PCR system (Applied Biosystems). Absolute quantification of IAV matrix 1 gene was done with standard plasmids and normalized by housekeeping gene GAPDH. Primers used in this study were:

GAPDH-F: GTCTCCTCTGACTTCAACAGCG;

GAPDH-R: ACCACCCTGTTGCTGTAGCCAA;

IAV-matrix gene-F: GGCATTTTGGACAAAKCGTCTA;

IAV-matrix gene -R: CTTCTAACCGAGGTCGAAACG

### Statistical analysis

The association of the monthly prevalence between all pairs of virus infections was tested using Spearman’s rank correlation coefficients. To assess whether IV prevalence was statistically associated with the evolution of the future values EV/RV prevalence, the Granger causality test was conducted and the significant lagged week of IV prevalence was determined.^18^ To examine the association between EV/RV and IV prevalence independent of meteorological effects at different lagged times, a quasi-Poisson generalized additive model (GAM) was used to control the total number of weekly collected samples (i.e. model offset), long-term trend, and seasonal trend. The technical detail has been noted in the Supplementary File. The effect of EV/RV on IV prevalence was quantified using adjusted relative risk (ARR) along with its corresponding 95% confidence interval (CI). The reference value was set as its median value.

The likelihood of viral co-infection was computed by Fisher’s exact test and logistic regression after adjustment to age and gender. Age group stratification with toddlers (age <2), preschool (age 2-5), school-age (age 6-17), adult (age 18-64) and elderly (age >65) were segregated for regression analysis. Differences in influenza titers and viral gene expression was compared at respective time points with or without prior EV exposure using two-way ANOVA followed by *Bonferroni* post-test for multiple comparisons. One sample t-test was used to compare the changes (log_10_ transformed) in IV titer with prior RV infection, with null hypothesis assuming no difference with sham treated control cells isolated from the same individuals. All statistical tests were performed using Graphpad version 9.2.0 and IBM SPSS Statistics. Differences were considered statistically significant at *p* < 0.05.

## Results

### Opposing seasonality of EV/RV and IVs

A total of 112,926 NPA were included in this study. EV/RV was the most prevalent viral infection (**Figure 1B**), and it contributed a monthly positive rate of at least 8% throughout the study period (**Figure 2**, red line). IVs were the second most dominant viral group being detected (**Figure 2**, navy line). Combining IAV, IBV and ICV, reached up to 35% positive rate during flu season but remained low for the rest of the year. A strong seasonal pattern was observed in both EV/RV and IVs, with robust biannual peaks of EV/RV occurred during spring and autumn, and one to two peaks of IV occurred in summers and winters of Hong Kong, yet the onset, magnitude, duration and dominating subtypes of the peaks varied extensively (**Figure 2**). A staggered pattern between EV/RV and IVs has been observed in which the intensity of flu peaks was often higher after a low EV/RV season. During the spring of 2017, the shortest period of EV/RV peak was followed by an early outburst of IAV of the H3N2 subtype.

**Figure 1.**
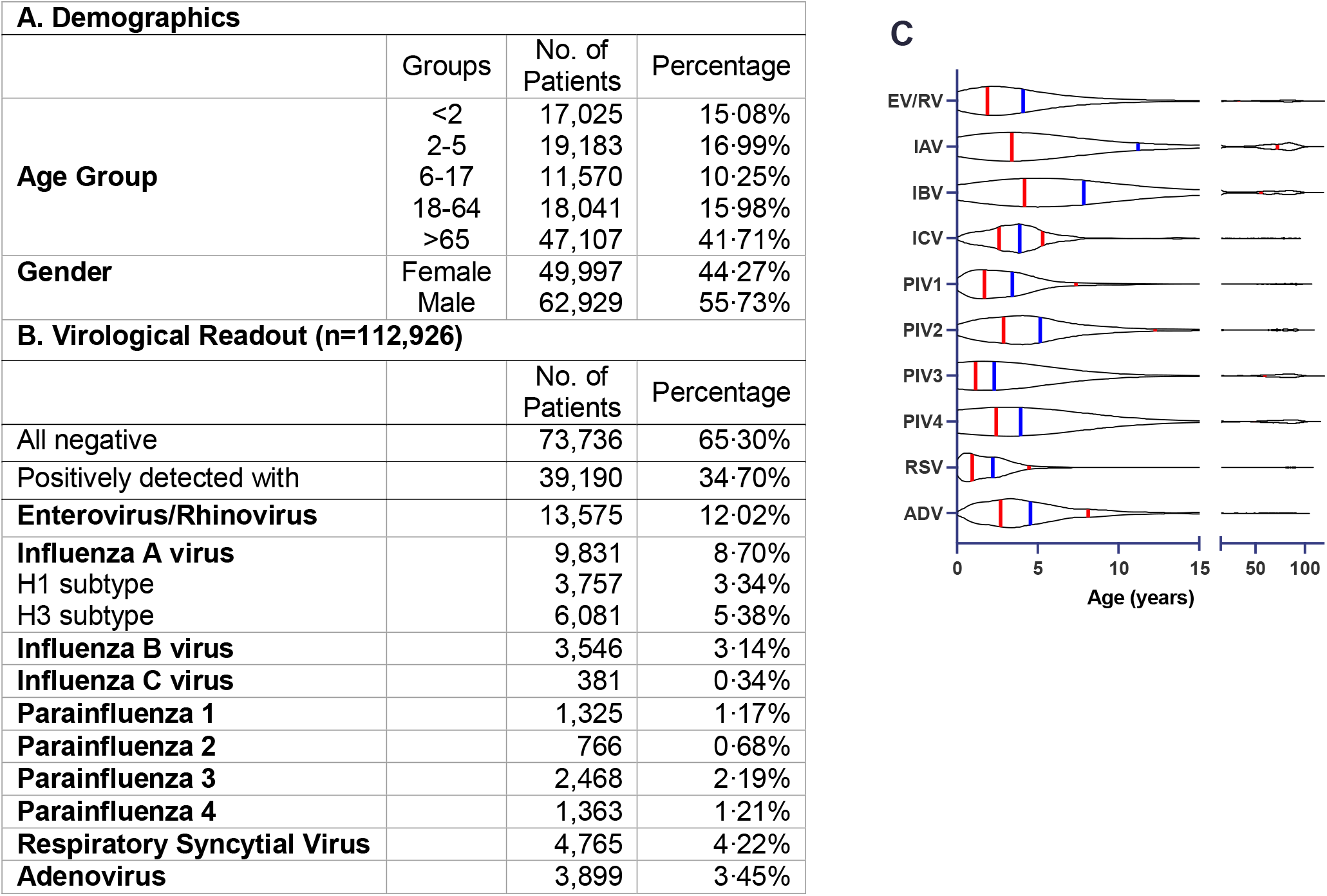
**(A)** Demographics of the patients included in the inpatient study from September 2015 to December 2019. **(B)** Virological readouts were obtained from all 112,926 NPA samples for the detection of virus infection by multiplex PCR. Subtyping of H1 and H3 were performed after positive IAV detection. **(C)** A violin plot showing the age distribution of virus infections from test-positive 39,190 NPA samples presents individual data as grey dots. The blue line shows the median age, and the red lines show the interquartile range.

**Figure 2.**
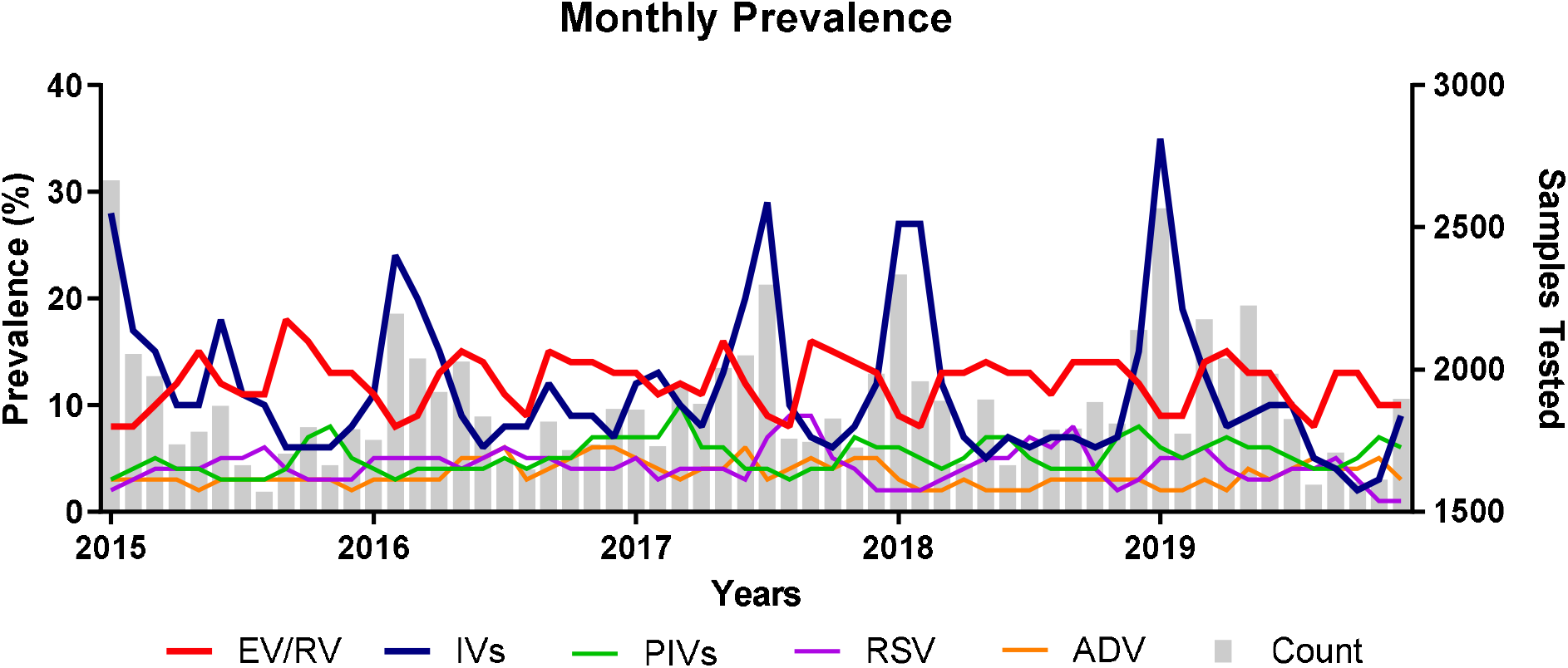
Temporal dynamics of respiratory virus prevalence in the inpatient cohort from September 2015 to December 2019. Monthly prevalence of individual virus infections (left y-axis) with the respective number of tested samples showed in grey bars (right y-axis) throughout the study period. EV/RV = enterovirus/rhinovirus; IVs = influenza viruses; PIVs = parainfluenza viruses; RSV = respiratory syncytial viruses; ADV =adenovirus.

### Negative correlation between the prevalence of EV/RV with IVs

Interactions between viruses may be confounded by other factors such as the age of the subjects and meteorological factors during sampling. In the current study, we analyzed the virological data by age stratification (**Figure 1C**). EV/RV was the most prevalent in those aged under 2 years old, while the median age of IAV and IBV positive cases was significantly higher (11.19 and 7.84 years old, respectively) than that of EV/RV (4.01 years old). Due to the variation of the influenza subtypes each year, IAV-H1, IAV-H3, IBV and ICV were combined as IVs for regression analysis. Logistic regression analysis revealed a significant negative correlation between the monthly prevalence of EV/RV against IVs (−0.421, *p<*0.01, **Figure 3C, navy**) but not with other virus pairs, such as PIVs, RSV and ADV. Spearman’s bivariate analysis showed a similar result in which a significant negative correlation was identified between EV/RV against total IVs or IAV (**Table 1**). Significant negative correlations between the monthly IVs prevalence and PIV2 and PIV4 were though their intensities were not as strong as that between IVs and EV/RV.

**Figure 3:**
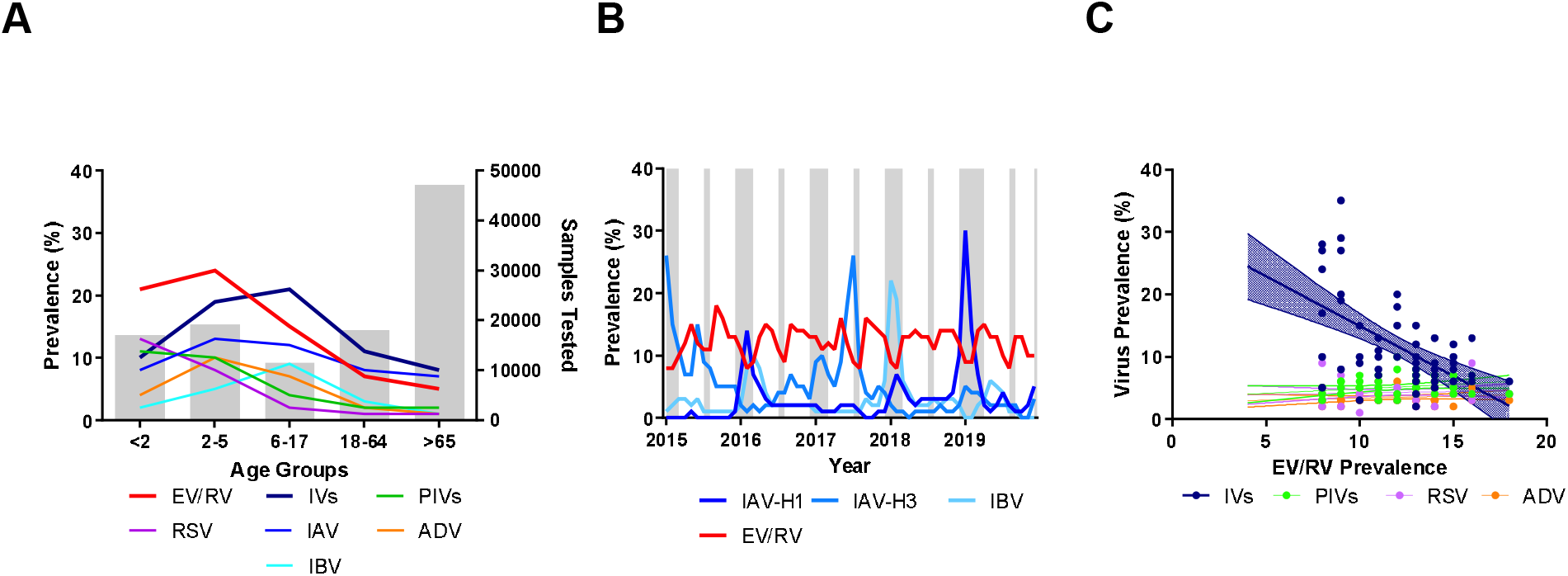
Negative correlation between the prevalence of EV/RV and IVs. **(A)** Prevalence of individual virus infections of different age groups is represented by the colour lines (left y-axis). The grey bar represents the number of samples tested in each group (right y-axis). **(B)** Prevalence of influenza subtypes (gradients of blue) compared to EV/RV (red) across the five-year study period. Typical influenza season (December to March, July to August) in Hong Kong is shaded in grey. **(C)** Logistic regression between the prevalence of EV/RV with other viruses with 95% CI marked in dotted lines. Significant negative correlation (−1.596 ± 0.3110, *p*<0.001^***^ with R^2^ = 0.3123) was identified only between EV/RV and IVs.

**Table 1.** Correlation of viral prevalence. Bivariate Spearman’s cross-correlation coefficients between respiratory viruses using the monthly prevalence are shown. Asterisks indicate significance at *p*<0.05^*^, *p*<0.01^**^ and *p*<0.001^***^. IVs in the last column indicates the sum of IAV, IBV and ICV, the cross-correlation within the IVs and IAV, IBV and ICV are therefore excluded from the analysis. Red and blue values indicate a negative and positive correlation, respectively.

| *p* | IAV | IBV | ICV | PIV1 | PIV2 | PIV3 | PIV4 | RSV | ADV | IVs |
| --- | --- | --- | --- | --- | --- | --- | --- | --- | --- | --- |
| EV/RV | -0.421** | 0.096 | -0.223 | 0.117 | 0.114 | 0.240 | 0.147 | 0.052 | 0.138 | -0.477*** |
| IAV |  | -0.217* | -0.025 | 0.126 | -0.350** | -0.184 | 0.042 | 0.078 | -0.120 | - |
| IBV |  |  | 0.081 | -0.186 | -0.289* | 0.224 | -0.232 | -0.022 | -0.100 | - |
| ICV |  |  |  | -0.147 | 0.277* | 0.138 | 0.025 | -0.426** | -0.125 | - |
| PIV1 |  |  |  |  | 0.038 | 0.275 | 0.415* | -0.172** | -0.051 | 0.047 |
| PIV2 |  |  |  |  |  | 0.098 | -0.053 | -0.178 | -0.090 | -0.428** |
| PIV3 |  |  |  |  |  |  | 0.084 | -0.396* | -0.011 | -0.061 |
| PIV4 |  |  |  |  |  |  |  | -0.047 | 0.059 | -0.109** |
| RSV |  |  |  |  |  |  |  |  | -0.032 | -0.044 |
| AdV |  |  |  |  |  |  |  |  |  | -0.199 |

### Time series causality

IV prevalence was significantly associated with the evolution of EV/RV (*p<*0.001) and the effect of IV was highly significant at lag zero (*p<*0.001), indicating a non-lagged interference between IV and EV/RV as assessed by the Granger causality test. The disease-disease association at lag zero was further examined via GAM analysis and a significant negative association between IV and EV/RV was showed (**Figure 4**). The ARR of EV/RV was 0.652 (95% CI: 0.571 to 0.745) when the prevalence of IV increased to 31.3% (i.e. 95^th^ percentile of IV), whereas the ARR of EV/RV was 1.159 (95% CI: 1.079 to 1.244) when the prevalence of IV decreased to 1.5% (i.e. 5^th^ percentile of IV), with a median reference value (9.3%).

**Figure 4.**
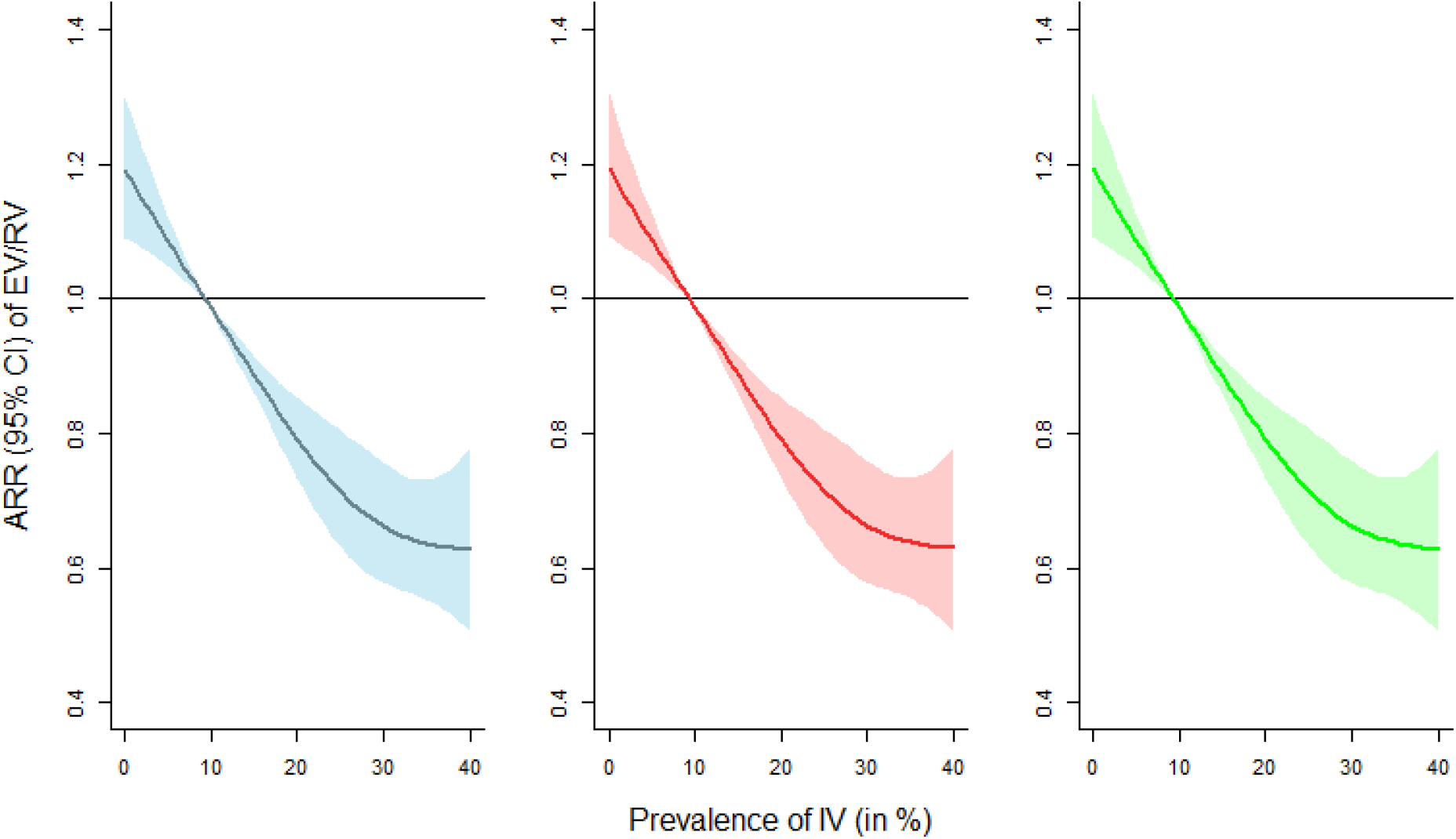
Adjusted relative risks (ARRs) with 95% confidence interval on EV/RV against IV prevalence. The estimated ARRs without controlling meteorological effects, with ambient temperature plus relative humidity controlled, and with actual vapor pressure adjusted are expressed as blue, red, and green colors respectively. Median IV prevalence was used as the reference value for comparison.

Moreover, the negative relationship between EV/RV and IV was independent to meteorological variations from both ambient temperature & relative humidity and absolute humidity. After controlling the effect of temperature and relative humidity, the ARR of EV/RV was 0.654 (95% CI: 0.572 to 0.748) when the prevalence of IV was at its 95^th^ percentile. An increase of lagged time of IV demonstrated a sinusoidal change in the ARR of EV/RV, highlighting a counteracting oscillation between the two infections (**Supplementary Figure 1**).

### Reduced likelihood of EV/RV and IVs co-infection

Co-infection of respiratory viruses is common in hospitalized patients. Overall, 9.1% (n=2,582) of the NPA samples were co-detected with two or more respiratory viruses, and 59.8% of these co-infection cases were contributed by EV/RV (n=1,545) (**Figure 5A**). Co-infections were more common in children, in which more than 80% of cases were found in paediatric patients with age under 18 **(Figure 5B)**. Interestingly, the odds to have both EV/RV-IVs detected in the same specimen was exceptionally low (OR=0.15) when compared with 0.75 for EV/RV-PIVs, 0.54 for EV/RV-RSV and 0.94 for EV/RV-ADV co-detection using Fisher’s Exact Test (**Figure 5C**). A further reduction in odds was observed between EV/RV and IVs after the adjustment of the confounding effect due to age and gender by binary logistic regression (**Figure 5D**).

**Figure 5.**
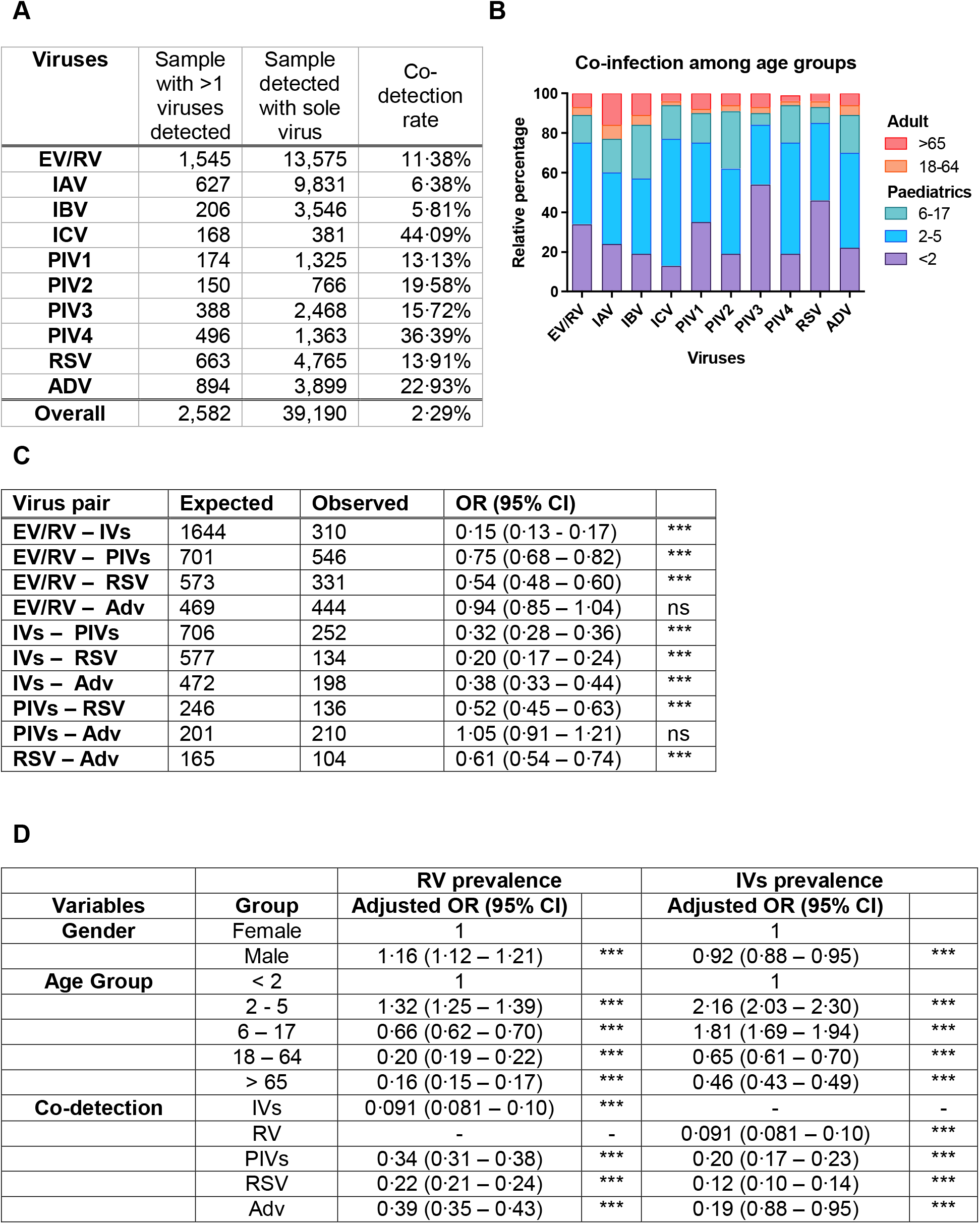
Co-infection statistics. **(A)** Rate of co-detecting more than one respiratory virus in different respiratory virus infection. The number of NPA samples detected with more than one pathogen detected was divided by the total number of the sample test-positive with the agent listed in each row. **(B)** Breakdown of co-infection cases according to age group. **(C)** The odd ratio of EV/RV and IVs co-infection with other respiratory pathogens using Fisher’s exact test with null hypothesis assuming the likelihood of individual infection events were not interrelated **(D)** Logistic regression analysis of EV/RV and IV infection after adjustment to gender and age group with the same adjustments.

### Sequential infection of RV and IV in HBEC and HNEC

To address if viral interference between IV and RV occurs at the cellular level, a sequential infection was performed on well-differentiated human airway epithelial cells (**Figure 6A**). RV-A16 and RV-1B and seasonal IAV, IBV and R-IAV showed productive replication and achieved a 2-log_10_ increase at 48 hpi. HBEC were susceptible to RV-A16 and RV-1B infection and replication without cytopathic changes, while IV infection led to obvious CPE at 48hpi (**Supplementary Figure 2**).

**Figure 6.**
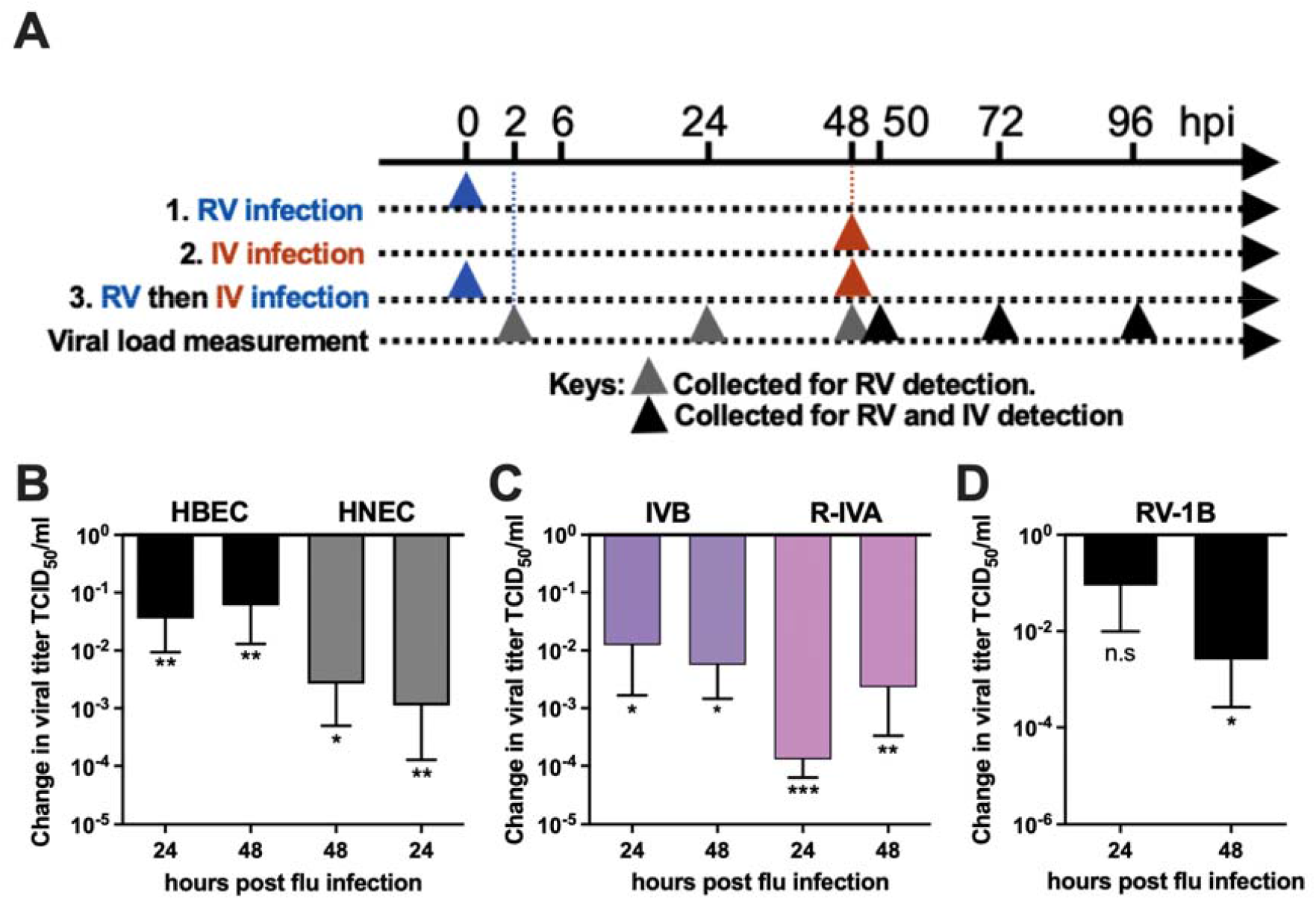
RVs suppress subsequent IVs infection in vitro. **(A)** Experimental plan of the model. Fully differentiated ALI cultures were first infected (or sham-treated) with RVs for 2 days, followed by secondary infection of IVs. Supernatant and cell lysates were collected at 2, 24 and 48 hours post infection (hpi) for determination of viral load and gene expression. **(B)** Suppression of IVA titers with prior RV-A16 infection in HBEC (n=9) and HNEC (n=5). Y-axis represents the difference in titer in RV exposed to the sham-treated cells. **(C)** Suppression of IVB and oseltamivir-resistance stain of IVA (R-IVA) in HBEC with prior RV-A16 exposure. **(D)** Suppression of seasonal IVA with prior RV-1B infection in HBEC. Error bars showing the SEM of means and asterisks indicating significance of *p*<0.05*, *p*<0.01** and *p*<0.001*** compared with sham treatment as examined by one-sample t-test.

### Suppression of IAV infection and replication with prior exposure to RV-A16 and RV-1B in HBEC and HNEC

Prior exposure to RV-A16, the IV replication was inhibited. The viral load was significantly inhibited with a mean reduction of 1.44 log_10_ and 1.22 log_10_ in HBECs and 2.95 log_10_ and 2.58 log_10_ in HNECs at 24 h and 48 h post influenza virus infection, respectively, compared with those exposed to sham treatment. (**Figure 6B**). A significant reduction in the normalized IAV matrix gene copies was also observed in both cell types (**Supplementary Figure 2I**).

### Active RV replication is essential to exert viral interference

Heterogenicity in individual’s susceptibility to RV was observed. HNECs derived from 4 out of 5 donors, and HBECs derived from 6 out of 9 donors supported productive replication of RV (**Supplementary Table 1 and Supplementary Figure 3)**. Importantly, we found that the interference effect depended on the active replication of RV. The inhibition effect to IAV infection was abolished in non RV-replicating cells. Using ultraviolet-inactivated RV-A16 as the inoculum, we confirmed this observation again and found that the UV-inactivated virus did not suppress the subsequent IVA and IVB replication.

### Suppression of IBV and oseltamivir-resistant IAV with prior exposure to RV-A16 and RV-1B in HBEC

The suppression by the prior exposure of RV-A16 was not limit to IAV. A significant reduction of IBV (1.91 log_10_, and 2.25 log_10_ at 24hpi and 48hpi, *p*<0.05) and oseltamivir-resistant IAV strain (3.85 log_10_ and 2.64 log_10_ at 24 and 48hpi respectively, *p*<0.01, **Figure 6C**) was observed in RV-A16 infected cells compared with sham-exposed HBECs. Moreover, RV exposure protected HBEC from IAV induced cell death. Extensive CPE was starting from 48hpi, while no CPE was observed for at least 7 days if the cells were infected with RV-A16 before IVA infection (**Supplementary Figure 2C to 2H**). To rule out if the observation is a specific effect of RV-A16, the same experiment setting was carried out using RV-1B in HBECs. RV-1B suppressed the IAV replication by 2.59 log_10_ at 48hpi but not at an earlier time point (**Figure 6D**). Taken together, our result supports that RV infection attenuates the subsequent influenza replication in primary human respiratory epithelial cells.

## Discussion

The influenza virus exerts a great burden on the health system each year in terms of frequent medical visits, hospitalization and flu-related death. We demonstrated the viral interference between RV and IVs using epidemiological data and biological experiments, suggesting a broad protective role of EV/RV in inhibiting subsequent IVs. We evaluated this interaction using the epidemiological data collected from hospitalized patients from September 2015, when the EV/RV test was first introduced as a routine test in the clinical settings, to December 2019, the last normal month before the SARS-CoV-2 pandemic began. In this study, a total of 112,926 NPA obtained from all ages were examined. The negative association between EV/RV and IVs prevalence was independent of subject age and meteorological factors. Consistent with studies performed in different climatic parameters, ^10,11^ an interference effect in population-level is suggested. We also demonstrated the competitive effect between the two could also operate at the individual level as the likelihood of getting co-detection between EV/RV and IVs was exceptionally low compared with other virus pairs. During these nine flu seasons in Hong Kong, it is intriguing to see that EV/RV prevalence oscillated in a counteracting manner. For example year 2015, 2018 and 2019 represented the lowered EV/RV positive rate of winters (November to February of the next year) within the study window were followed by some of the most intense winter flu outbreaks. The opposite is true for the summer of 2018. While experiencing the highest summer EV/RV prevalence within the period, the intensity of flu within the same season was almost abolished. The magnitude of concurrent RV peak may forecast the intensity of IVs in the same season which is beneficial for public health management before flu outbreaks.

To convey the observation of viral interference from *in vitro* settings to the population level, a transmission study using animal models will be a choice. It has been shown that prior exposure to RV-1B can reduce the severity of mouse-adapted IAV PR8 in a dose-dependent manner. ^19^ In the same study, mice with a double-stranded RNA (dsRNA) mimic before IAV infection significantly reduce viral load. Aligned with our findings in cells that did not support RV replication and UV-inactivated RV, it has been shown that UV-inactivated RV cannot induce antiviral cytokine responses.^20^ The lack of inhibitory effect to subsequent IAV infection suggested that active virus replication within the host is required. Viral interference may be mediated by factors such as IFNs, defective interfering particles, production of trans-acting proteases, cellular factors, and nonspecific dsRNA.^21^ A recent finding from an *in vivo* mouse model suggested that inhibition of IAV PR8 by RV depended on type I IFN signalling pathway.^22^

It has been shown that nonpharmaceutical interventions such as ethanol hand rub and facemasks are not effective in controlling the transmission of non-enveloped EV/RVs ^23,24^. During the SARS-CoV pandemic, hospitalization and positive rate for detected enveloped virus including IVs, PIVs and RSV were drastically reduced but that of EV/RV was less affected throughout 2020 ^25^. Excluding the period of SARS-CoV-2 pandemic is necessary to address the interference effect in the community as the implementation of social distancing measures and enhanced personal hygiene has a great effect in suppressing the transmissibility of respiratory viruses. Using a similar experimental design, our preliminary *in vitro* infection model suggested that prior RV-A16 infection could suppress SARS-CoV-2 replication using HBEC (**Supplementary Figure 4**). A recent finding suggested that the protective effect conferred by prior EV exposure in respiratory cells is again due to the induction on IFN-stimulated genes shared high consistency with our findings ^26^.

*In vitro* infection using ALI differentiated cells and mathematical simulations also agree that RV has an interference effect against SARS-CoV-2 at multiple levels. ^27^ These results all point to the fact that EV/RV infection, which usually causes mild or asymptotic infection in healthy individuals, may provide at least transient protective effect against more virulent viral infections such as influenza and SARS-CoV-2. Immunomodulatory effect due to mild EV/RV effect may serve as a novel antiviral defense against emerging outbreaks when therapeutics are not available. Understanding the molecular mechanism on how EV/RV triggering innate immunity may shed light on novel prophylactics design against board range of viral infections.

### Limitations

The viral interference window induced by RVs to IVs *in vitro* was up to 48 hours and the maximal duration is yet to define. We demonstrated that the inhibitory effect was significant for at least 48 hours post influenza virus infection. However, the exact durability of the inhibition exerted by RV was not thoroughly assessed in this setting because of extensive cell death in the sham-treated cells **(Supplementary Figure 3H)**. Moreover, we did not investigate if the suppression of IAV by RV relate to impairment of influenza virus receptor. Nevertheless, a previous study showed that RV-A16 infection on HBEC did not alter the transcription of α2-6-linked and α2-3-linked sialyl-transferases, which are responsible producing the relevant sialic acid receptor, thus the restriction on IVA is independent of the change in receptor availability.^28^ Lastly, the action of competition between RV and IV by adding the virus together at the same time point or prior infection of IV to subsequent RV infection was not evaluated. The latter could not be evaluated in our human primary respiratory epithelial cell culture, as the IV would cause a significant CPE even at a low MOI.

## Conclusion

The cumulative evidence suggests the occurrence of viral interference at the population, individual and cellular level. The understudied role of RV in providing the baseline immunity to influenza virus replication warrants further attention.

## Data Availability

All data in the manuscript is available upon request.

## Acknowledgements

We would like to acknowledge Professor Gillian M Air of the University of Oklahoma Health Sciences Center who provides IAV strain A/Oklahoma/447/2008 H1N1 for the study; Drs Steffi X Long and Louisa LY Chan for culturing the human primary epithelial cells the study; Ms Waii WY Yu for her assistance in the molecular biology experiments. Drs Michael CW Chan and Dr Kenrie PY Hui for the SARS-CoV-2 infection.

## Authors contributions

Conception, experimental design, drafting the manuscript and interpretation: KPT, MKCC and RWYC; Data Analysis: KPT, MKCC, JSCP, MHW, RWYC; Collection of bio-specimen: JGST, SMWC, CSHN, ZC, PKSC, AML; Execution of experiment and acquisition of data: KPT and JCSP. All authors reviewed and approved the final manuscript.

## Notes

### Competing Interest Statement

The authors have declared no competing interest.

### Author Declarations

This study was approved by the Joint Chinese University of Hong Kong New Territories East Cluster Clinical Research Ethics Committee (CREC: 2015.097 and 2019.120).

