## Supplementary Appendix for "Suppression of influenza virus infection by rhinovirus interference at the population, individual and cellular levels"

| **Table A: Subjects who donated their HNEC** \| \| \| \|; **Patient no.** \| **Gender** \| **Age** \| **Complaints and diagnosis** \| **RV Replication**; 467 \| M \| 30-35 \| Nasal obstruction with rhinitis symptoms, acute pharyngitis, hypertrophy of nasal turbinates and deviated nasal septum \| Y; 469 \| F \| 35-40 \| Nasal swelling \| Y; 470 \| M \| 30-35 \| Hypertrophy of nasal turbinates, bilateral turbinate reduction, allergic rhinitis with obstructive symptoms \| Y; 477 \| M \| 60-65 \| Previous history of lung adenocarcinoma, later confirmed to have cancer of hypopharynx with EBV >80 and VCA >320 \| N; 478 \| M \| 55-60 \| Hypotrophy of nasal turbinates, rhinitis with nasal obstruction \| Y **Table B. Subjects who donated their HBEC.** | | | | |
| --- | --- | --- | --- | --- |
| **Patient no.** | **Gender** | **Age** | **Surgical site and principal diagnosis** | **RV Replication** |
| 37 | F | 70-75 | Right middle lobe with adenocarcinoma | Y |
| 38 | F | 60-65 | Left lower lobe nodule with adenocarcinoma with predominant acinar pattern | N |
| 39 | M | 85-90 | Left upper lobe adenocarcinoma | Y |
| 41 | F | 60-65 | Left upper lobe with benign nodules | Y |
| 42 | M | 70-75 | Right upper lobe adenocarcinoma with predominant acinar pattern | N |
| 45 | F | 70-75 | Left lower lobe moderately differentiated adenocarcinoma and GGO nodule | N |
| 46 | F | 65-70 | Left lower lobe non-small cell adenocarcinoma and lymphoepithelioma-like carcinoma | Y |
| 47 | F | 65-70 | Right lower lung moderately differentiated adenocarcinoma with predominant acinar pattern | Y |
| 56 | F | 70-75 | Right upper lobe moderately differentiated adenocarcinoma with predominantly lepidic pattern | Y |

**Supplementary Table 1:** Demographics of patients for **(A)** HNEC and **(B)** HBEC isolation.

Productive RV replication was defined as ≥ 0.5 log_10_ increase in TCID_50_/ml after 48 hours at 0.01 MOI when compared to 2 hours post infection. Specific age is replaced by age range to remove identifying information. Patient no. showed is not an identifiers that can reveal the identity of the study subjects outside the research group.

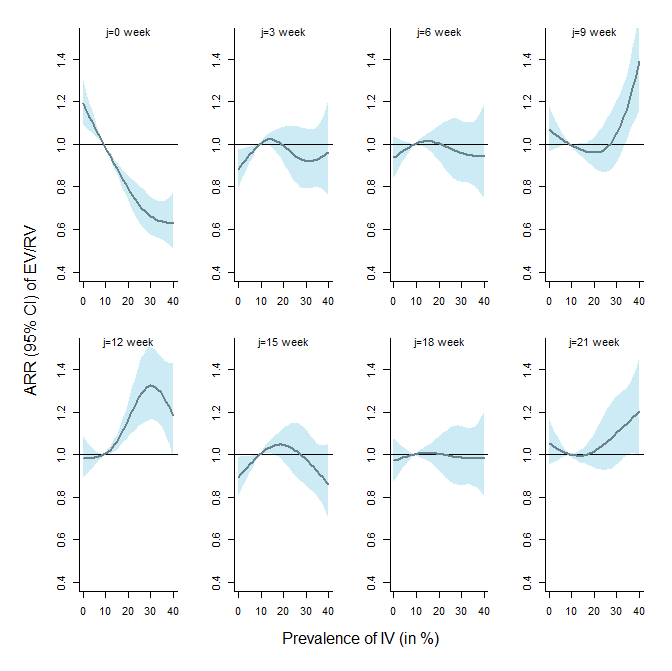

**Supplementary Figure 1. Adjusted relative risks (ARRs) with 95% confidence interval on EV/RV against IV prevalence at different lagged times.** The estimated ARRs without controlling meteorological effects were obtained via a GAM analysis setting with different lagged weeks (*j*) of IV prevalence (i.e. *IV_t-j_*). Median IV prevalence was used as the reference value for comparison.

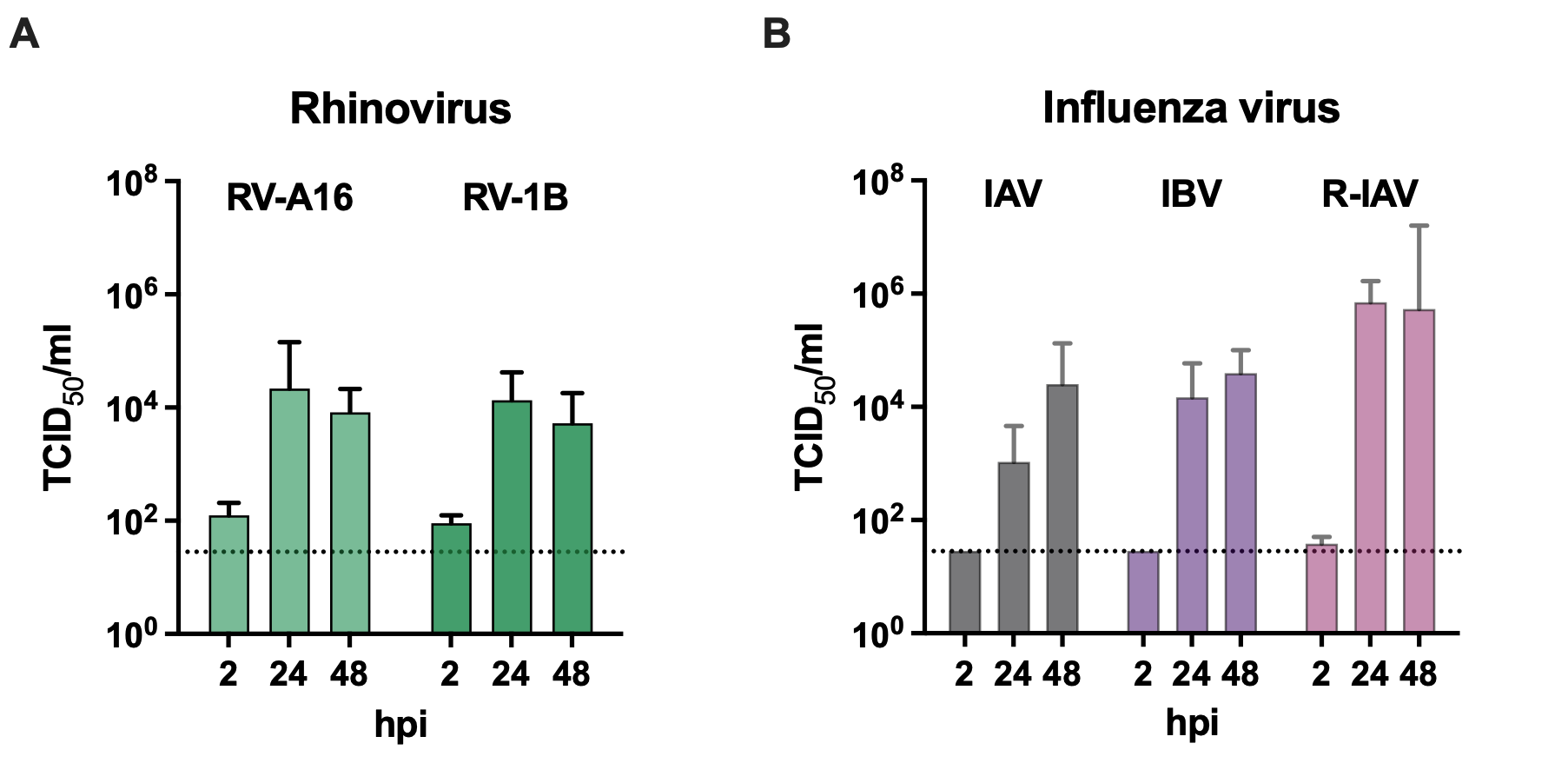

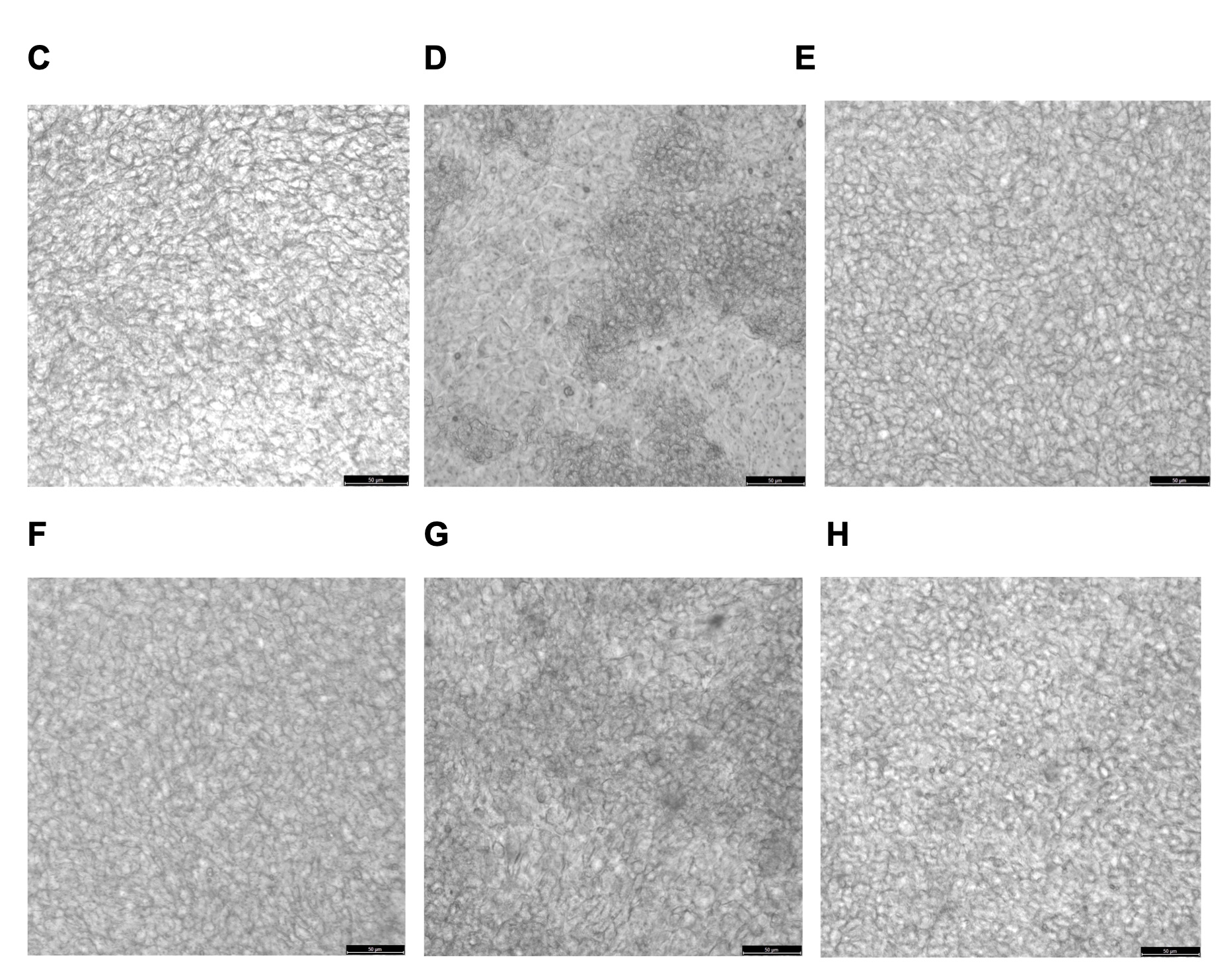

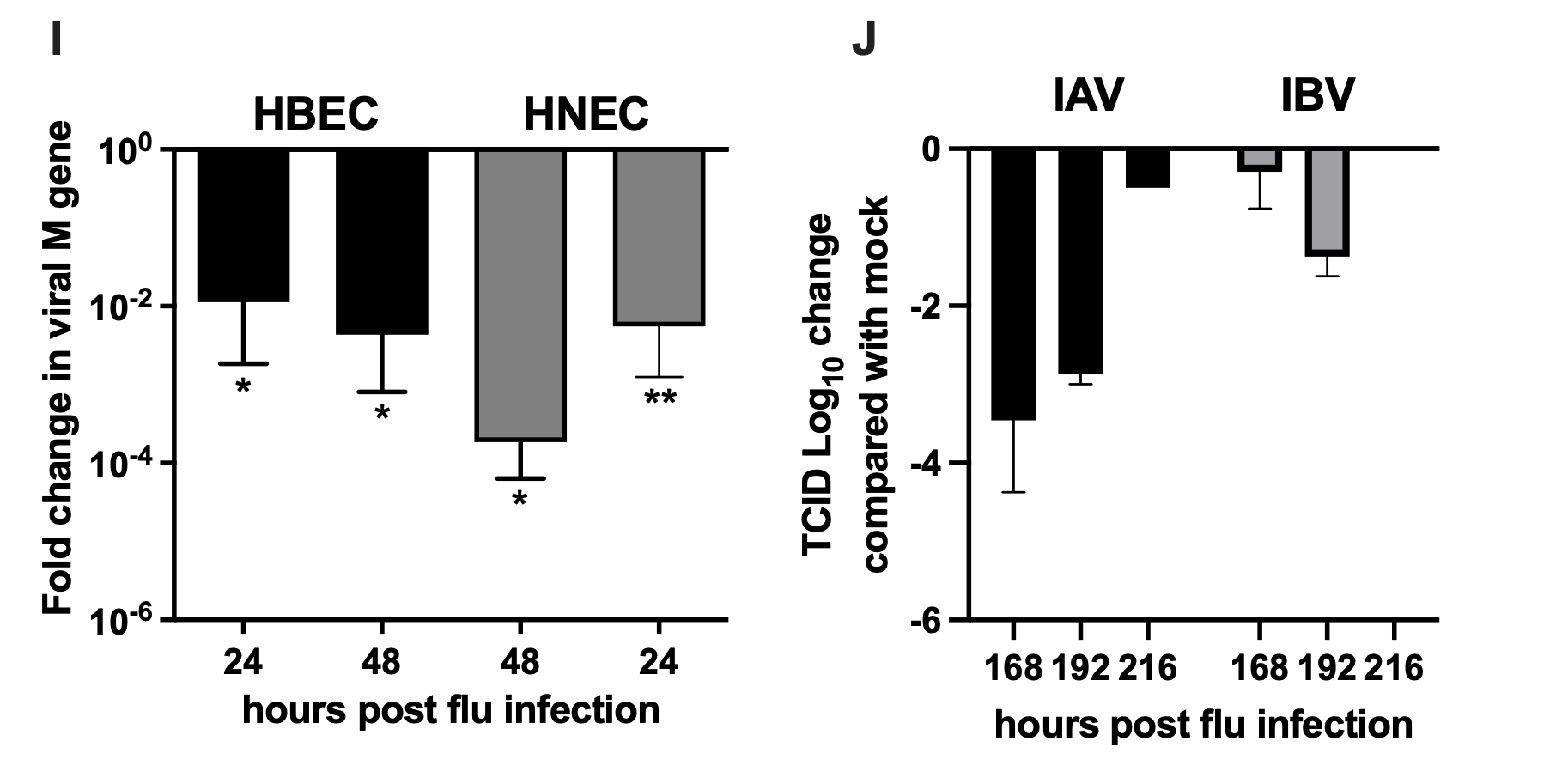

**Supplementary Figure 2.** Viral load of **(A)** RV-A16 and RV-1B and **(B)** IVs from the supernatant in HBEC at the first 48 hour time points as measured by viral titration assay and expressed in TCID_50_/ml. Dotted lines represented the detection limit of the assay. Cytopathic effect in HBEC after **(C)** Mock treated, **(D)** IAV only, **(E)** RV-A16 then IAV **(F)** RV-1B then IAV **(G)** IBV only, and **(H)** RV-A16 then IBV. Images were acquired after 48 hours of primary infection followed by 48 hours of secondary infection. Scale bar = 50 um. **(I)** Suppression of IV matrix gene mRNA copies in HBEC and HNEC by qPCR. **(J)** Interference effect in extended time points. Reduction of flu titers upon RV-A16 pre-treatment was identified at 144, 168 and 192 post IVA infection compared to sham control. Experiments were conducted in selected donors only (n=2) due to extensive cell death upon IVA infection at 96hpi.

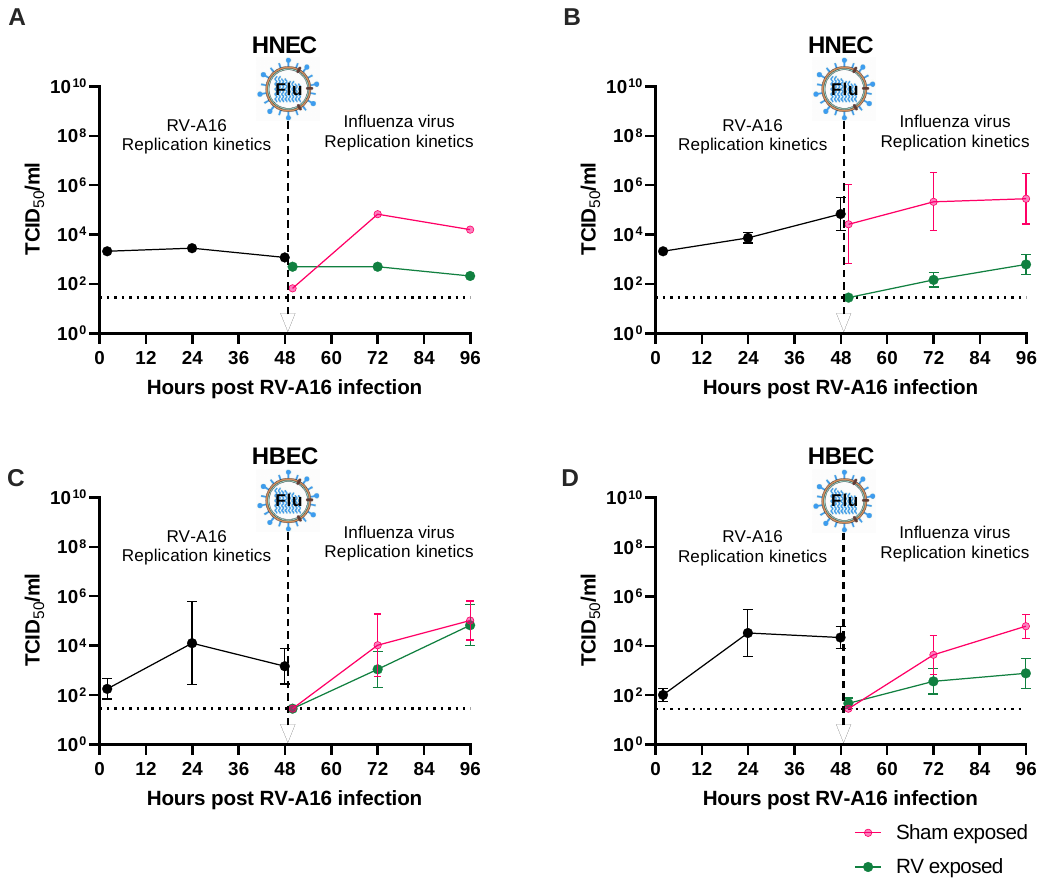

**Supplementary Figure 3. IAV interference on HNEC and HBEC depends on productive replication of RV.** Productive replication of RV was defined as ≥ 0.5 log_10_ increase in virus titer upon the infection of RV-A16 at an MOI of 0.01. **(A and C)** HNEC (patient 477) and HBEC (patients 38, 42 and 45) with ineffective RV-A16 replication in the first 48 hours resulted in no IAV suppression effect. **(B and D)** HNEC and HBEC with productive replication of RV-A16 in the first 48 hours exerted significant IAV suppression (green line) when compared to the sham exposed cell (sham exposed).

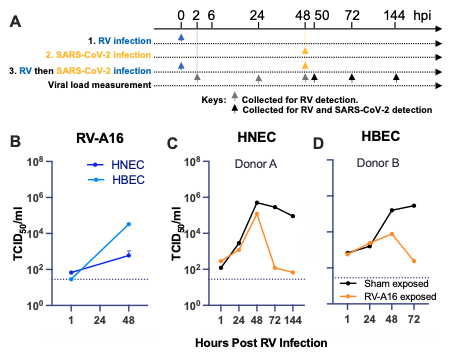

**Supplementary Figure 4. Experimental design of the sequential RV-A16 and SARS-CoV-2 virus infection and the RV inhibition effect on SARS-CoV-2 in well-differentiated HNECs and HBECs.** SARS-CoV-2 from a patients with confirmed COVID cases were isolated and propagated in Vero E6 cells (1). Infection experiments were performed in biosafety level 3 facility at School of Public Health, Li Ka Shing Faculty of Medicine, The University of Hong Kong. Primary HNECs on transwell inserts were exposed to **1)** RV-A16, **2)** SARS-CoV-2 at a multiplicity of infection (MOI) of 0.01 and 1, respectively; or **3)** a prior RV infection at MOI = 0.01 then a SARS-CoV-2 at 48 hpi of the initial RV inoculation. SARS-CoV-2 virus was inoculated to the same culture for 1 h followed by washing steps to remove the inoculum. The supernatant from the apical compartment of the transwell inserts were collected at 1, 24, 48, 72 and 144 hpi by adding 200ul PBS for the detection of the **(B)** RV-A16 viral load detected in HNEC (navy) and HBEC (cyan) in the first 48 hours. SARS-CoV-2 viral load expressed in 50% tissue culture infectious dose (TCID_50_)/ml in **(C)** HNEC and **(D)** HBEC (black). Cells that were prior infected with RV-A16 (orange) are susceptible to SARS-CoV-2 infection at the first 48h for HNEC and 24h for HBEC followed by a suppression of viral titer of 4 log_10_ and 2 log_10_ at 72hpi and 48 hpi, respectively. The suppression continued to the last time point of supernatant collection, i.e. 144hpi and 72 hpi, respectively.

**Supplementary note. Technical details of Granger causality test and generalised additive modelling.**

**Granger causality test**. Granger causality is commonly used in the field of econometric analysis. We employed this test to assess whether the information of IV was able to generate an improvement in the prediction of EV/RV. Under this framework, the null hypothesis was set as IV did not Granger-cause EV/RV,

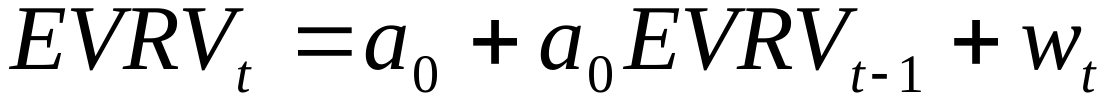

where *EVRV_t_* is the EV/RV prevalence at time *t*, *a_0_* is the model intercept, and *a_1_* is the model coefficient quantifying the effect of EV/RV prevalence at different lagged time i.e. *t*-1, *t*-2, …. The term *w_t_* is the model residual at time t and assumed to follow a normal distribution with mean zero and a standard deviation of σ. The alternative hypothesis is thus set as IV Granger-cause EV/RV,

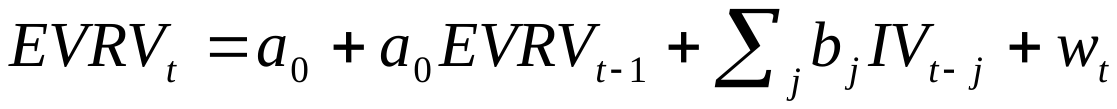

where *IV_t-j_* is the IV prevalence at *j*^th^ lagged week and *b_j_* is the model coefficient quantifying the corresponding lagged effect of IV prevalence.

**Generalised additive modelling**. We employed quasi-Poisson generalized additive models to examine the non-linear disease-disease association between EV/RV and IV controlling long-term trend and seasonal trend. The model form is as follow

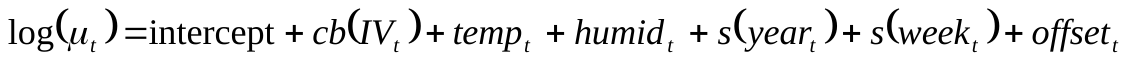

where *μ_t_* is the expected number of EV/RV cases and *offset_t_* is the natural logarithms of the total number of weekly collected samples on week *t*. The function *cb*(.) denotes a cross-basis function and *s*(.) represents a smoothing spline function with degree of freedom equal to 4 as a usual setting. The terms *year_t_* and *week_t_* were included in the model to capture long-term trend and seasonal trend respectively. Weekly mean temperature (*temp_t_*) and relative humidity (*humid_t_*) were controlled in the model. To examine the association controlling with actual vapor pressure (*e*), the two meteorological terms in the model were replaced with it,

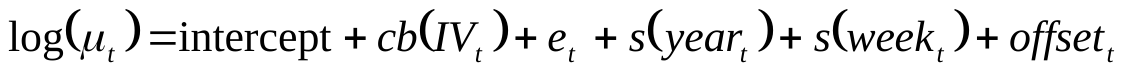

where *e_t_* is the actual vapour pressure on week *t*.
